# A global perspective of the prevalence of low language in children: a scoping review and evidence and gap map protocol

**DOI:** 10.1101/2024.07.25.24310917

**Authors:** Lisa Irene Jones, Michelle A Krahe, Amy Giesberts, Martin Downes, Robert S Ware, Sheena Reilly

## Abstract

**Objective:** The objective of this scoping review is to identify, categorize, and map the existing evidence of low language prevalence in children aged 4 to 18 years globally. Additionally, it seeks to explore associated factors, and map evidence gaps to improve future reporting practices.

**Introduction:** The study of language difficulties in children has evolved over the years, marked by changing terminologies and criteria. Understanding why prevalence varies is critical to address the need for accurate prevalence estimates, highlight factors influencing reported frequency, and inform policy, resource allocation, and research priorities.

**Inclusion criteria:** This review will consider studies that report low language prevalence for children aged 4 to 18 years from any country that are derived from studies or epidemiological samples in community/population settings, including households, birth registers/cohorts, and schools. To encompass the changing use of terminologies and criteria, the term low language is used as an umbrella term to describe children whose language development is behind their peers.

**Methods:** This review protocol is guided by the Joanna Briggs Institute methodology for scoping reviews. Key databases will be systematically searched for English-language studies, published between 01 January 1970 to 30 June 2023. Two independent reviewers will screen titles and abstracts, while three reviewers will assess full texts. The final selection of studies will undergo review by a broader expert group and data extraction will be conducted by one reviewer and verified for accuracy. The findings will be presented in narrative format, including tables and figures to aid in data presentation.

## Introduction

Whilst not all children learn language at the same rate, it is well recognized that a group of children have problems acquiring language. The first descriptions of children having such difficulties appeared nearly two centuries ago(1), and since that time there has been a proliferation of terms for language difficulties, with some used briefly, whereas others have gained greater traction(2). A pivotal debate in 2014 prompted an international consensus panel to examine the terminology and criteria used to describe language problems in children(3, 4). This resulted in the endorsement of the term Language Disorder (LD) to reflect “… *a profile of difficulties that causes functional impairment in everyday life and is associated with poor prognosis*”(5, p1068) and the term Developmental Language Disorder (DLD) to describe children who have difficulties learning language in the absence of any known biomedical etiology(5, 6).

The lack of agreement about the terminology and criteria used to identify children with language problems has been described as detrimental to research(5) including our understanding of the prevalence of the condition. Estimates remain imprecise, ranging from 7-17% between cohort studies(7-9). In a recent systematic review, Hill, Calder (10) reported even wider estimates, from 0.4% to 25.2% in children aged 1 to 18 years, with greater stability of language level observed in children aged 5 years and older. We note that these findings are not specific to the study of childhood language; similar variation in prevalence has been observed in other developmental conditions including autism, attention-deficit/hyperactivity disorder (ADHD). Further, reports demonstrated that some of the factors that influenced prevalence estimates, included the child age at which prevalence was estimated, the measure/s used, the definition and diagnostic criteria of the disorder, and the geographic location and population of the sample(11).

Accurate prevalence estimates are essential for informing public policy, raising awareness, and developing research priorities(12). They are instrumental in resource allocation, influencing funding decisions, and the expansion of service provision. Therefore, understanding why current prevalence estimates vary is important. There are likely to be multiple factors that influence estimates including the study design, the definition and measurement of language problems, and the age of the child at assessment. As in the study of other developmental conditions, the geographical location may also play a role. Identifying these factors could influence precision in current reporting practices and inform future research directions. A global estimation of prevalence serves as a valuable tool for advocacy and guides research focus, particularly in regions where prevalence data is limited or unavailable(13).

At the outset we recognized that this review would need to consider the changing terminology and criteria used to describe children with language problems in the search strategy. After Hill, Calder (10) we adopted the term ‘low language’ to ensure we captured a wide range of reports on language ability in children. The authors limited the terms they used to describe the number of children with language problems to prevalence, incidence, and epidemiology and may have inadvertently missed published studies that either did not use these terms or where reports of low language prevalence were embedded in broader published reports, such as in nationally representative birth cohorts. We therefore adopted broad search criteria to capture published reports from a range of studies that might have commenced at any time over the past 40-50 years, during which terminology and criteria will have changed.

We anticipate encountering numerous studies reporting on the prevalence or incidence of low language worldwide as well as variability in the study contexts, including the type of sampling and its derivation, as well as variation in how low language was determined. This encompasses considerations such as when and how measurements were taken, including the age of the child at measurement, the tools used for measurement, the language domains measured and the reporting methods employed (e.g., total scores, subgroup scores). Furthermore, we anticipate encountering studies reporting on comorbidities associated with low language.

To our knowledge, there appear to be no formal reviews of factors associated with the prevalence of low language in children, globally. The objective of this review is to provide an overview of the evidence related to low language prevalence in children, and to identify, map and categorize factors associated with prevalence reporting. The findings from this review will have several practical implications. Firstly, it will provide a comprehensive understanding of the global prevalence of low language in children aged 4 to 18 years, thereby informing policymakers, healthcare professionals, and educators about the scope of this issue. Secondly, by exploring factors driving the prevalence within the studies, such as population age, measurement methods, and geographic location, the study can identify gaps in the current literature and inform low language prevalence reporting in future studies. Finally, mapping evidence gaps will help identify areas where further research is needed, facilitating the improvement of reporting practices and ultimately leading to more accurate prevalence estimates and better-informed decision-making in healthcare and education sectors.

## Review question(s)

1. What are the key characteristics of studies that report prevalence of low language in children aged 4 to 18 years?
2. What are the factors associated with the reporting of prevalence?
3. What are the gaps in the current literature, and how do they, and the findings of this scoping review, inform low language prevalence reporting in future studies?

## Inclusion criteria

### Participants

This review will consider studies that include children aged 4 to 18 years old(14). Four years was selected as the lowest age limit because language development in the early years is characterized by greater fluctuation and stabilizes by 4 years of age(15, 16).

### Concept

This review will consider studies that describe or evaluate the prevalence of low language, which is used to describe a group of children whose language development is behind their peers. This can include terms such as developmental language disorder, specific language impairment or language delay. This concept search was designed using key terminology for language disorders and guided by expert researchers within the field. Low language is determined by a specific language measure(s) such as standardized language assessment tools, observational methods (i.e., language samples collected during naturalistic interactions), parent or teacher reported measures, or a combination of multiple measures that provide a comprehensive evaluation of language abilities. Language outcome determined by report only, clinical referral, or ascertained from a clinical record will be out of scope.

### Context

This review will consider studies where data is derived from studies or epidemiological samples in community or population settings, including households, birth registers/cohorts, and schools. This includes cross-sectional, prevalence, incidence, cohort, demographic, proportion, epidemiology, or population studies. Excluded will be studies involving clinical, referred or volunteer-based samples, and samples assembled retrospectively. Studies that lack sufficient information to estimate prevalence or those devoid of original data, such as literature reviews or commentaries, will also be excluded from consideration.

### Types of sources

This review will consider studies that have been conducted in any geographic location or setting. Both experimental and observational study designs, including randomized controlled trials, non-randomized controlled trials, before and after studies, and cohort or population studies will be considered.

## Methods

The proposed scoping review will be conducted in accordance with the Joanna Briggs Institute (JBI) methodology for scoping reviews(17, 18). The review will report findings following the Preferred Reporting Items for Systematic Reviews and Meta-Analyses extension for Scoping Reviews (PRISMA-ScR)(19). This protocol has been registered with the Open Science Framework (https://osf.io/mkns2).

### Search strategy

The search strategy will aim to locate both published primary studies, and reports of primary studies. An initial limited search of MEDLINE (via Ovid) was undertaken to identify articles on the topic. The text words contained in the titles and abstracts of relevant articles, and the index terms used to describe the articles, were used to develop a full search strategy for MEDLINE (via Ovid) in Appendix I. Collaboration with a member of the Griffith University Library ensured the accuracy and comprehensiveness of the search strategy, which will be adapted for each included information source. The final search strategy will be run on various databases, including MEDLINE via Ovid, ERIC via ProQuest, Psycinfo via Ovid, Web of Science, Scopus and CINAHL via EbscoHost and the reference lists of articles selected for inclusion will be screened for additional papers. Only articles published in English between 01 January 1970 to 30 June 2023 will be included in the final review.

### Study selection

Following the search, all identified records will be collated and uploaded into the review management software Covidence (Veritas Health Innovation, Melbourne, Australia). The software will automatically remove duplicates, and this will be checked to ensure accuracy. Following an initial pilot test, two independent reviewers will screen titles and abstracts to assess their alignment with the inclusion criteria. Potentially relevant papers will undergo full retrieval, imported into Covidence, and assessed in detail against the inclusion criteria by three independent reviewers. Any full-text papers not meeting the criteria will be recorded and reasons will be included in the scoping review. Disagreements among reviewers will be resolved through discussion. The final selection of studies will undergo review by a broader expert group. The search results will be presented in the final scoping review and depicted in a flow diagram following the Preferred Reporting Items for Systematic Reviews and Meta-Analyses extension for Scoping Reviews (PRISMA-ScR) guidelines(19, 20).

### Data extraction

Data will be extracted by one reviewer using a data extraction tool developed by the reviewers. The data extraction tool will be tested independently by three reviewers on three studies to ensure that specific details about the population, concept, context, and study methods of significance, to ensure consistency in extraction. The draft data extraction tool is provided in Appendix II and will be modified and revised as necessary during the extracting process. Modifications will be detailed in the full scoping review. Any disagreements that arise between the reviewers will be resolved through discussion. Authors of papers will be contacted to request missing or additional data, where required.

### Data analysis and presentation

Studies included in this scoping review will be analyzed using quantitative descriptive analysis and will be visualized through evidence and gap maps. The characteristics of studies reporting the prevalence of low language in children, the types of samples involved (i.e., countries, cohorts, language disorder), along with their respective study design and key language outcomes will be described. Additionally, this review will provide an analysis of factors associated with prevalence, categorizing them as evidence for positive, negative, or neutral influence. Potential variations in the definitions of low language across studies will undergo quantitative content analysis, utilizing inductive categories. A bibliographical analysis will be used to conduct content analysis of the current research evidence, capturing prominent research themes in the field, and identifying evidence gaps. Knowledge translation activities will include publication in a peer-reviewed journal and conference presentations. Through direct collaboration with global, national and local networks including research collaborations and partnerships, professional associations, clinical groups, organizations responsible for policy setting and resource allocation and through organizations for people with language disorder and their families.

## Data Availability

All data produced in the present work are contained in the manuscript

## Acknowledgements

We would like to thank the Early Language in Victoria Study (ELVS) team of investigators, past and present, who have provided input.

## Author contributions

LIJ, MAK, AG & SR contributed to the initial conceptualization, drafting and finalization of this manuscript. MD and RSW made substantial contributions to the finalization of the protocol. All authors have reviewed and approved the complete manuscript.

### Appendix I: Search strategy

#### MEDLINE (Ovid)

Search conducted on 3 July 2023

| PCC framework | # | Keywords and Medical Subject Headings (MESH) terms | Records retrieved |
| --- | --- | --- | --- |
| Population:<br>Children<br>aged 4-18<br>years old | 1 | (exp Young Adult/ OR exp Birth Cohort/ OR exp Child/<br>OR exp Child, Preschool/ OR exp Adolescent/) | 3,816,086 |
|  | 2 | limit 1 to yr="1970 -Current" | 3,605,150 |
|  | 3 | ("young adult*" or "birth cohort" or child* or preschool* or<br>pre-school* or kindergarten* or youth or adolescen* or<br>teen*).ti,ab. | 1,723,065 |
|  | 4 | limit 3 to yr="1970 -Current" | 1,641,013 |
|  | 5 | 2 OR 4 | 3,944,424 |
| Concept:<br>Prevalence<br>of low<br>language | 6 | Language Development Disorders/ or Specific<br>Language Disorder/ | 7,331 |
|  | 7 | limit 6 to yr="1970 -Current" | 7,297 |
|  | 8 | ("language development disorder*" or "specific language<br>disorder*" or "specific language impairment*" or<br>"developmental language disorder*" or "expressive<br>receptive language disorder*" or "primary language<br>impairment*" or "language learning impairment*" or<br>"language delay*" or "language disorder*" or "language<br>impairment*" or "speech and language impairment*" or<br>"low language" or "primary language impairment*").ti,ab. | 8,273 |
|  | 9 | limit 8 to yr="1970 -Current" | 8,209 |
|  | 10 | 7 OR 9 | 12,464 |
| Context:<br>Type of<br>study | 11 | exp Cross-Sectional Studies/ or exp Prevalence/ or exp<br>Incidence/ or exp Cohort Studies/ | 3,223,996 |
|  | 12 | limit 11 to yr="1970 -Current" | 3,203,696 |
|  | 13 | ("cross sectional stud*" or "cross-sectional stud*" or<br>prevalence or "prevalence stud*" or incidence or<br>"incidence stud*" or "cohort stud*" or "demographic stud*" or rate* or proportion* or epidemiolog* or<br>"population stud*").ti,ab. | 4,862,091 |
|  | 14 | limit 13 to yr="1970 -Current" | 4,823,384 |
|  | 15 | 12 OR 14 | 6,379,905 |
| Final Search | 16 | 5 AND 10 AND 15 | 3,354 |

### Appendix II: Draft data extraction instrument

| Category | Data to be extracted |
| --- | --- |
| Article information | • Author(s) and year of publication<br>• Year language data collected<br>• Study location |
| Study design | • Aim(s) of the study<br>• Population and sampling (type and how derived) |
| Participant characteristics | • Age at recruitment (range, mean, and SD)<br>• Gender (distribution/ratio)<br>• Socioeconomic status<br>• Comorbidity examined |
| Study stage and sample | • Sample size<br>• Sample characteristics, including how sample was ascertained (inclusion/exclusion criteria if applicable) |
| Assessment details | • Age at assessment<br>• Screening/assessment method (how/which measures administered)<br>• Diagnostic tool (and/or criteria) and how outcome on tool/criteria was reported (i.e., total score, subscale score, subgroups)<br>• Language domain assessed in the measures (i.e., vocabulary [expressive/receptive], pragmatics, narrative, semantics, syntax, morphology, omnibus, composite) |
| Low language identification | • Terminology used to identify language disorder (i.e., SLI, DLD, language difficulty)<br>• How disordered language is determined (i.e., school data, clinical assessment, score on measure)<br>• Prevalence of language difficulty as specified |

## References

1. McKean C, Law J, Morgan A, Reilly S. Developmental language disorder. United Kingdom: Oxford University Press; 2018.

2. Law J, Reilly S, McKean C. Language Development: Individual Differences in a Social Context: Cambridge University Press; 2022.

3. Reilly S, Bishop DV, Tomblin B. Terminological debate over language impairment in children: forward movement and sticking points. International Journal of Language & Communication Disorders. 2014;49(4):452–62.

4. Bishop DVM. Ten questions about terminology for children with unexplained language problems. International Journal of Language & Communication Disorders. 2014;49(4):381–415.

5. Bishop DVM, Snowling MJ, Thompson PA, Greenhalgh T, consortium C. CATALISE: A Multinational and Multidisciplinary Delphi Consensus Study. Identifying Language Impairments in Children. PLOS ONE. 2016;11(7):e0158753.

6. Bishop DVM, Snowling MJ, Thompson PA, Greenhalgh T. Phase 2 of CATALISE: a multinational and multidisciplinary Delphi consensus study of problems with language development: Terminology. Journal of Child Psychology and Psychiatry. 2017;58(10):1068–80.

7. Tomblin JB, Records NL, Buckwalter P, Zhang X, Smith E, O’Brien M. Prevalence of specific language impairment in kindergarten children. Journal of Speech, Language, and Hearing Research. 1997;40(6):1245–60.

8. Reilly S, Wake M, Ukoumunne OC, Bavin E, Prior M, Cini E, et al. Predicting language outcomes at 4 years of age: findings from Early Language in Victoria Study. Pediatrics. 2010;126(6):e1530–7.

9. Norbury CF, Gooch D, Wray C, Baird G, Charman T, Simonoff E, et al. The impact of nonverbal ability on prevalence and clinical presentation of language disorder: evidence from a population study. Journal of Child Psychology and Psychiatry. 2016;57(11):1247–57.

10. Hill E, Calder S, Candy C, Truscott G, Kaur J, Savage B, et al. Low language capacity in childhood: A systematic review of prevalence estimates. International Journal of Language & Communication Disorders. 2024;59(1):124–42.

11. Le HND, Le LKD, Nguyen PK, Mudiyanselage SB, Eadie P, Mensah F, et al. Health-related quality of life, service utilization and costs of low language: A systematic review. International Journal of Language & Communication Disorders. 2020;55(1):3–25.

12. Zeidan J, Fombonne E, Scorah J, Ibrahim A, Durkin MS, Saxena S, et al. Global prevalence of autism: A systematic review update. Autism Res. 2022;15(5):778–90.

13. Checchi F, Warsame A, Treacy-Wong V, Polonsky J, van Ommeren M, Prudhon C. Public health information in crisis-affected populations: a review of methods and their use for advocacy and action. The Lancet. 2017;390(10109):2297–313.

14. United Nations Children’s Fund. The Convention on the Rights of the Child: The children’s version [Available from: https://www.unicef.org/child-rights-convention/convention-text-childrens-version#:~:text=A%20child%20is%20any%20person,under%20the%20age%20of%2018.

15. Hayiou-Thomas ME, Dale PS, Plomin R. Language impairment from 4 to 12 years: prediction and etiology. J Speech Lang Hear Res. 2014;57(3):850–64.

16. Ukoumunne OC, Wake M, Carlin J, Bavin EL, Lum J, Skeat J, et al. Profiles of language development in pre-school children: a longitudinal latent class analysis of data from the Early Language in Victoria Study. Child Care Health Dev. 2012;38(3):341–9.

17. Peters MDJ, Marnie C, Tricco AC, Pollock D, Munn Z, Alexander L, et al. Updated methodological guidance for the conduct of scoping reviews. JBI Evid Synth. 2020;18(10):2119–26.

18. Peters M, Godfrey C, Khalil H, McInerney P, Soares C, Parker D. 2017 Guidance for the Conduct of JBI Scoping Reviews. Joanna Briggs Institute Reviewer’s Manual 2017.

19. Tricco AC, Lillie E, Zarin W, O’Brien KK, Colquhoun H, Levac D, et al. PRISMA Extension for Scoping Reviews (PRISMA-ScR): Checklist and Explanation. Annals of Internal Medicine. 2018;169(7):467–73.

20. Page MJ, Moher D, Bossuyt PM, Boutron I, Hoffmann TC, Mulrow CD, et al. PRISMA 2020 explanation and elaboration: updated guidance and exemplars for reporting systematic reviews. BMJ. 2021;372:n160.

